# Evaluation of Clinical Outcomes of Riluzole monotherapy and Riluzole based adjunctive interventions in Amyotrophic Lateral Sclerosis: A meta-analytic and unsupervised clustering approach

**DOI:** 10.64898/2026.02.24.26346710

**Authors:** Hirtik Singh Rathore, Jashandeep Singh Brar, Srishti Gupta, Nancy Dalla, Saarthak Kumar, Hardik Singh Rathore, Dibyajyoti Banerjee, Subodh Kumar

## Abstract

Amyotrophic Lateral Sclerosis (Lou Gehrig’s disease) is a progressive neurodegenerative disease affecting hundreds of thousands of people worldwide. It is characterized by the degeneration of the neurons in the brain and spinal cord of the patients, leading to a loss of control of muscles. Over time, without nerves to stimulate them muscles tend to atrophy. ALS may occur sporadically or run in families; many mutations have been identified for the latter. Treatment of ALS is mostly limited to three approved therapeutic agents: riluzole, edaravone, and tauroursidiol/ sodium phenylbutyrate. Among these, riluzole remains the most effective despite its early discovery. There are no conclusive meta-analysis comparing riluzole monotherapy to all possible co-therapies present. In this work we have attempted to address such a concern and observed that no adjunct therapy significantly improved the performance of riluzole. However, mitochondrial/ oxidative stress modulator and neuroimmune/ neuroexcitability modulator co-therapy exhibited positive trends. Surprisingly, trials were mainly confined to the USA and European countries, indicating unequal demographic representation in ASL research. We have concluded that large double blinded inter-continental RCTs to be carried out for better understanding of the scenario.

## Introduction

Amyotrophic lateral sclerosis (ALS) is a neurodegenerative disease challenging the motor capabilities of the individual [1]. The death of nerve cells in the central nervous system leads to motor abnormalities as the muscles atrophy[1], leading to the characteristic degeneration associated with ALS. The number of ALS-affected people is around 0.26 to 23.46 per 100,000 people per year worldwide, with the median survival of patients after diagnosis being 2 to 5 years [2][3]. There are two categories of ALS, i.e., familial ALS and sporadic ALS. There are various known ALS-causing mutations, including SOD1, C9orf72, TARDBP, FUS, etc., which result in familial ALS [4][5][6][1]. While most cases are sporadic in nature without a known cause, there is an interplay of genetic and environmental factors. However, the pathogenesis of ALS shares many features irrespective of cause.

With the variation in onset and progression of symptoms in ALS, accurate measurement of disease progression remains a challenge. Due to the lack of well-established biomarkers, disease progression is measured in terms of the revised Amyotrophic Lateral Sclerosis Functional Rating Scale (ALSFRS-R) score [7)][8].The scale ranges from 0 to 48 and assesses four functional domains, each weighted equally: bulbar, gross motor, fine motor, and respiratory [9]. The primary system affected in ALS is the nervous system, wherein both types of motor neurons, i.e., upper and lower, are degenerated; it also significantly affects other systems, particularly the muscular system, in which muscle cells weaken and ultimately undergo atrophy. Resultantly, the digestive system is also affected in bulbar functions, as are the respiratory muscles. So, while measuring the progression of ALS, all aforementioned factors must be considered. Symptoms of these systems are components of the ALSFRS-R scores [10][11]. While considering the modalities, not many treatment strategies are available. There is no known treatment capable of fully stopping and reversing the condition. The most potent, efficacious standard care of treatment, riluzole, is used in ALS management and was approved in 1995 [12]. It is a glutamate blocker that reduces the excitation by glutamate, a plausible mechanism of nerve degeneration. Riluzole has a significant positive effect on the decline and survival of ALS patients [13]. The next drug to come into use in 2017 was edaravone, a free radical scavenger, which prevents neuronal death due to oxidative stress [14]. It exhibited a moderate decline in progression but lacked consistent benefits. So, the idea of using it as an add-on to riluzole was carried forward [15]. Further trials faced the same problem. None of the drugs approved so far has been able to surpass riluzole in efficacy or prolongation of life. This study aims to assess whether the present co-therapies render any benefit to ALS patients compared to using riluzole as monotherapy, which is of prime importance for public health.

## Methods

### Inclusion Criteria

1. Randomised, placebo-controlled trials
2. Patient Population: Diagnosed with Amyotrophic Lateral Sclerosis (ALS) – familial or sporadic, or a mixture of both
3. Intervention: Riluzole was given to all participants along with treatment intervention or a placebo.

### Exclusion Criteria

1. Non-human studies, including animal studies and preclinical studies
2. Non-randomised, placebo-controlled trials
3. Traditional medicine with unspecified/complex composition
4. Studies with incomplete/unavailable datasets

### Database Search

A literature search was conducted across five key databases up to September 2025, including Google Scholar, Embase, Web of Science, SCOPUS, and PubMed. The following keywords were optimised for best search results in each database: **PubMed**- (“Amyotrophic Lateral Sclerosis” [Mesh] OR ALS OR “Lou Gehrig Disease” OR “Motor Neuron Disease”) AND (“Riluzole” [Mesh] OR riluzole) **Google Scholar** - Was manually searched after the results from these keywords (‘amyotrophic lateral sclerosis’ OR ‘lou gehrig disease’ OR ‘motor neuron disease’) AND ‘riluzole’ AND (‘randomized controlled trial’ OR ‘randomized controlled trial’ OR ‘randomized controlled trial’) **Embase and Web of Science** - (amyotrophic+lateral+sclerosis OR lou+gehrig+disease OR motor+neuron+disease) AND (riluzole) AND (randomized+controlled+trial OR randomised+controlled+trial) **Scopus**- (amyotrophic+lateral+sclerosis OR lou+gehrig+disease OR motor+neuron+disease) AND (riluzole) AND (randomized+controlled+trial OR randomised+controlled+trial) AND (“amyotrophic lateral sclerosis” OR ALS OR “Lou Gehrig’s disease” OR “Lou Gehrig disease” OR “Lou Gehrig’s syndrome” OR “Lou Gehrig syndrome”) AND (riluzole OR (riluzole AND (“add-on” OR “add on” OR adjunct OR adjunctive OR “combination therapy”)))

### Screening of the articles

After a keyword-based search, the studies were downloaded in the RIS file format and uploaded to Rayyan2025. Duplicates were identified using Rayyan2025 and manually removed. Abstract-based filtering was done. The resulting articles were procured as full texts, and the articles with incomplete or unavailable data were excluded. The resulting pool of articles was subjected to full-text screening on the basis of exclusion and inclusion criteria, and any conflict between authors was addressed during the process. Articles that were unanimously agreed upon were included in the study, the PRISMA for the same can be referred at ***Fig.1***.

**Fig.1.**
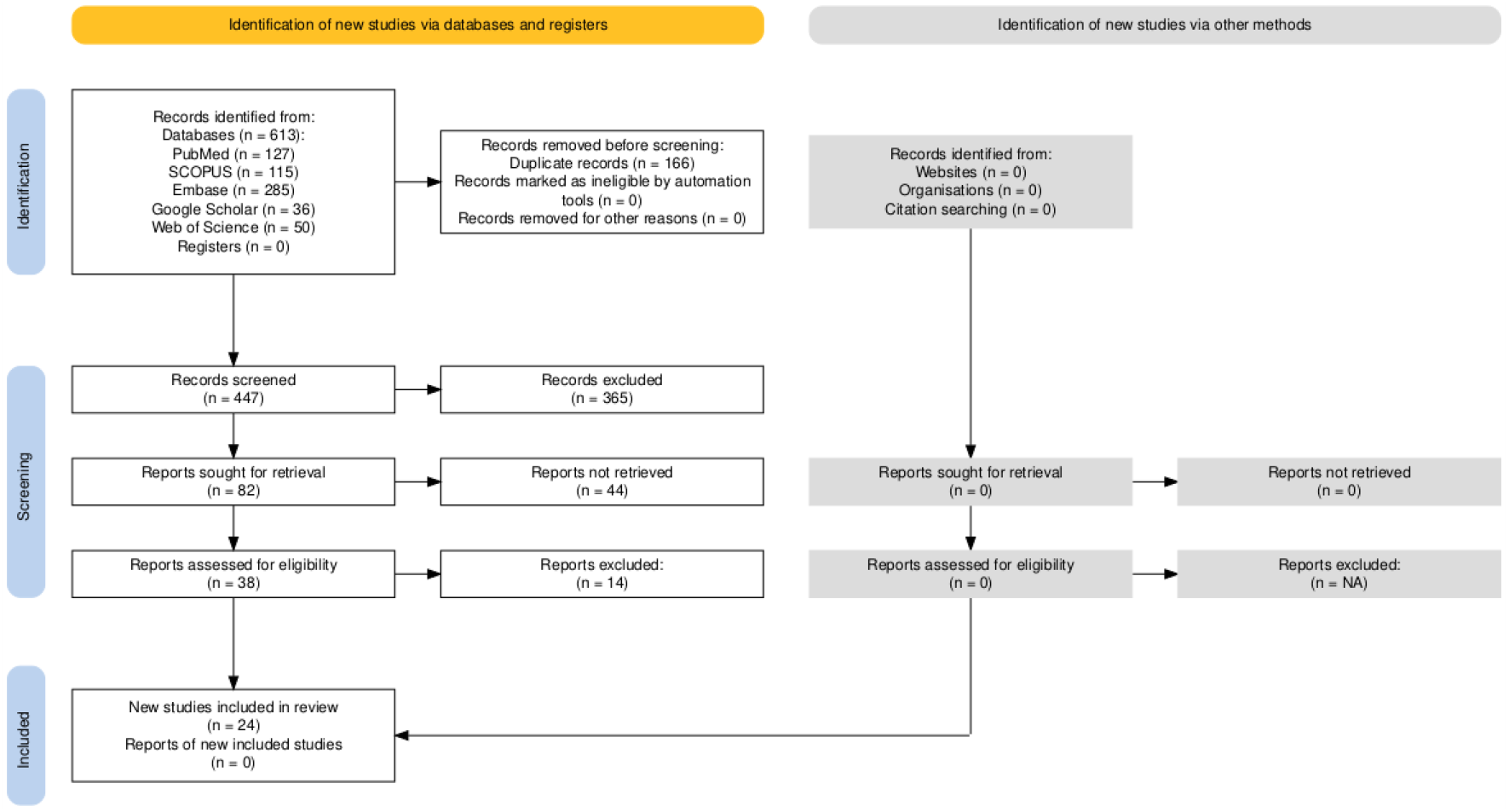
PRISMA flow Chart demonstrating the identification, screening, retrieval, eligibility, and inclusion of the studies in the meta-analysis.

### Data extraction and analysis

The parameters reported in the included studies were analysed. Data from common parameters was manually added in Microsoft® Excel® for Microsoft 365 MSO (Version 2510 Build 19328.20158). The efficacy of co-therapies was evaluated based on the change in the ALSFRS-R score and survival.

Given the degenerative nature of the disease, the difference in pre-treatment and post-treatment ALSFRS-R scores provides a good measure of decline in patient health. For the studies that reported a change in ALSFRS-R scores, most were reported as mean (standard deviation). For the studies that reported other measures of central tendency, the mean and standard deviation were estimated using established methods. Least squares mean was reported as such, and the standard deviation was calculated from the standard error. Mean and standard deviation were calculated from median and class interval. Survival, defined as “living without death, tracheostomy, or permanent assisted ventilation”, was analysed as risk ratios calculated from the number of surviving participants in both treatment and placebo groups. The safety of the drugs in combination with riluzole was evaluated based on the odds ratios of adverse events in treatment and placebo. The analysis was done using the R software (version 4.3.2; R Core Team, 2024). Thereafter, the demographics of the studies were also taken into consideration with the countries and years in which the studies were published to assess the gap in the literature with plausible hints at the ongoing research in the field of therapeutics in ALS.

### Risk of Bias and Publication Bias

The risk of bias was evaluated on the basis of bias from the randomization in the study, bias attributing to the deviations from the intended interventions, bias due to outcome data which is missing, and the bias in measurement of the outcome. All done using Cochrane Risk of Bias (RoB2). Additionally, publication bias was also evaluated using the funnel plot with statistical analysis.

## Results

### Survival and endpoint outcome analysis for ALS patients on therapy

As seen in ***Fig.2***, the cohort of all co-therapies with riluzole, only guanabenz at various doses exhibited statistically significant results with narrow confidence intervals, wherein riluzole monotherapy has better survival than co-therapy. Other interventions showed extended confidence intervals and non-significant results, while hinting at improved survival in some interventions, like acetyl-L-carnitine and arimoclomol. Dimethyl fumarate, growth hormone, and high-dose vitamin E favoured riluzole monotherapy over respective co-therapy. Moreover, mexiletine depicted no difference between both groups. On analysis with a random effects model, the overall pooled estimate indicates no significant difference between monotherapy and co-therapy. Heterogeneity was moderate (1^2^ = 39.3%).

**Fig.2.**
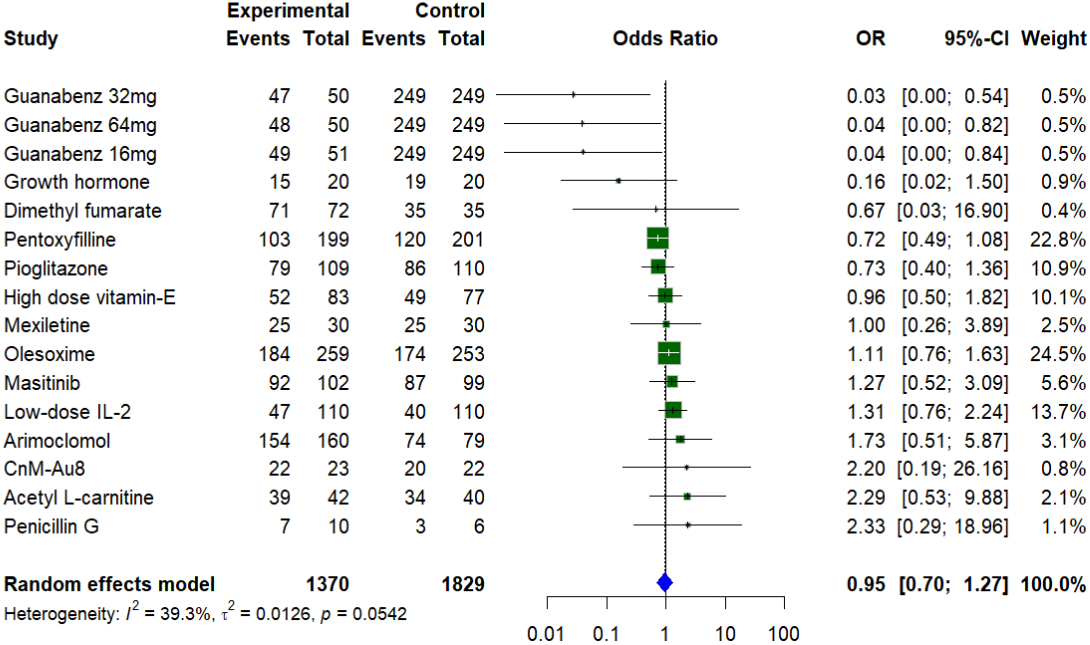
Forest plot of odds ratios for survival in cotherapy (experimental treatment) vs. monotherapy (control) in ALS clinical trials. Each square represents the odds ratio (OR) for survival in cotherapy vs. monotherapy, with the size of the square proportional to its weight in the meta-analysis. Horizontal lines indicate 95% confidence intervals (CI), and the diamond represents the pooled odds ratio under a random-effects model. The overall pooled estimate indicates no significant difference between experimental and control groups (OR = 0.95, 95% CI [0.70, 1.27], p = 0.0542). Heterogeneity was moderate (I^2^ = 39.3%)

### Condition improvement in patients on the basis of ALSFRS-R

In ***Fig.3(a)*** This meta-analysis of multiple studies comparing various treatments to placebo in ALS patients shows a pooled mean difference (MD) of 0.07 (95% CI: -0.29 to 0.43) in monthly ALSFRS-R score changes, indicating a very slight but statistically non-significant slowing of functional decline with treatment. Individual therapies such as CnM-Au8, Penicillin G, and Rasagiline demonstrated positive trends toward clinical benefit, whereas others like Arimoclomol and Dimethyl fumarate showed negative or minimal effects. The high heterogeneity observed (I^2^ = 92.2%) reflects considerable variability among study results, likely due to differences in study design, sample sizes, and treatment modalities. Overall, these findings suggest no conclusive evidence that adjunct treatments significantly improve ALS progression compared to placebo, highlighting the need for larger, well-controlled trials to better assess the efficacy of individual therapies. In ***Fig.3(b)*** Cerbrolysin in this analysis indicate a positive effect on slowing ALSFRS-R decline compared to placebo. However, it comes from a small study with only 10 patients in the treatment group and 8 in placebo, resulting in a low weight in the overall meta-analysis. While the result appears promising, the limited sample size means this finding should be interpreted cautiously and needs confirmation in larger, well-powered trials.

**Fig.3.**
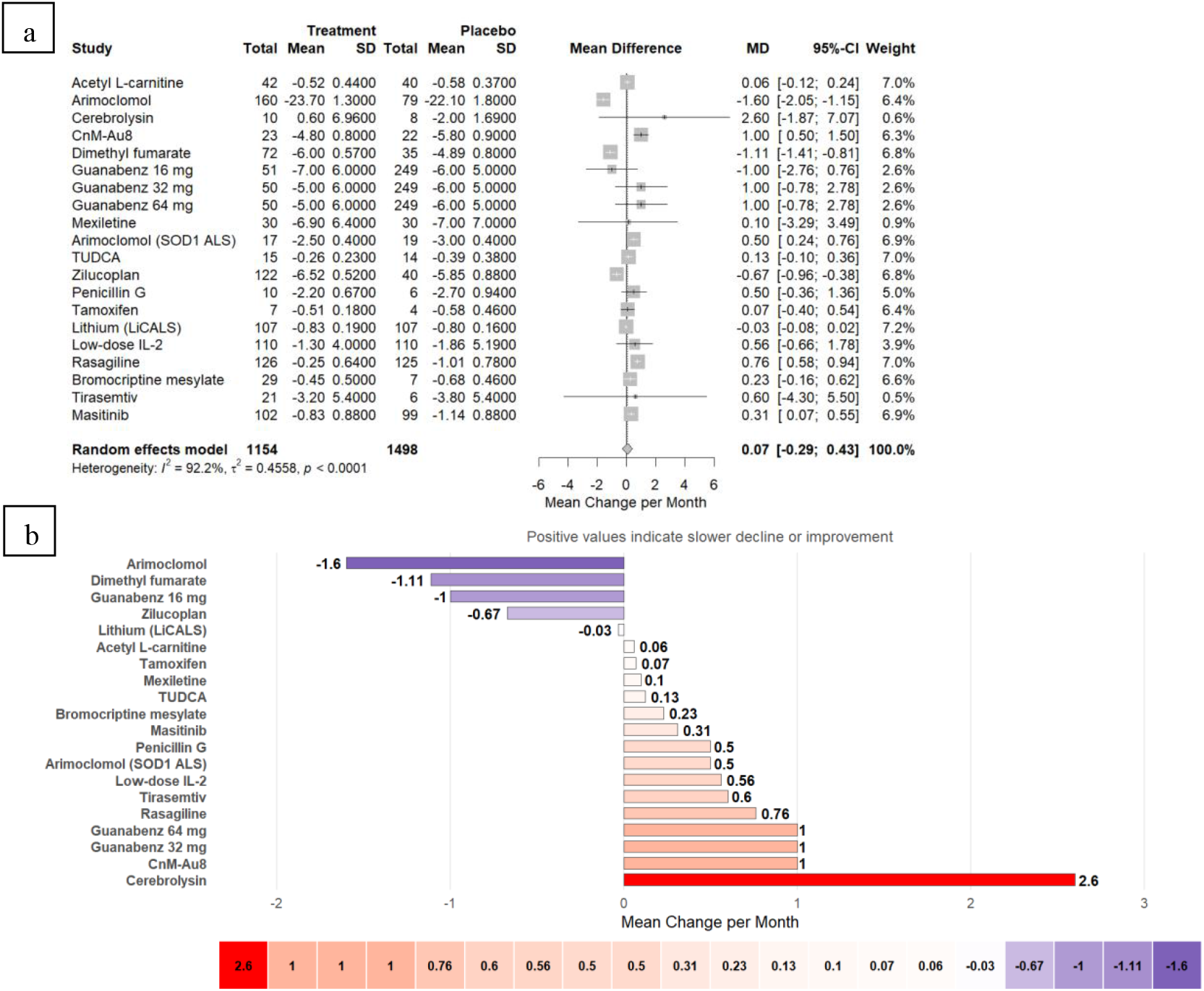
***(a)Forest plot of mean change per month in ALSFRS-R scores for treatment versus placebo groups.*** Each square represents the mean difference (MD) between treatment and placebo for an individual study, with the size of the square proportional to its statistical weight in the random-effects meta-analysis. Horizontal lines indicate 95% confidence intervals (CI). The diamond represents the pooled mean difference across studies (MD = 0.07, 95% CI [-0.29, 0.43], p < 0.0001). The positive pooled estimate indicates a slower functional decline in the treatment group compared to placebo. Heterogeneity was substantial (I^2^ = 92.2%). (***b) Comparison of mean change per month in ALSFRS-R scores across treatment groups***. This bar plot displays the mean change per month in ALSFRS-R scores for each treatment compared to placebo, with positive values indicating slower decline or clinical improvement. Treatments associated with greater improvement (e.g., Cerebrolysin and Guanabenz) are shown in red, while those showing greater decline or less benefit (e.g., Arimoclomol, Dimethyl fumarate) are shown in purple. The color gradient reflects the direction and magnitude of the mean change, facilitating visual comparison of therapeutic effects across studies.

### Evaluation of Publication Bias and Risk of Bias

As seen in ***Fig.4(a)*** the funnel plots for both the survival/endpoint and ALSFRS-R outcomes exhibited symmetry in the distribution of effect size around the pooled estimate, suggesting lack of publication bias. The analysis was augmented statistically with Egger’s Test and Begg’s Test scores (survival analysis [Egger’s Test = -1.35, p = 0.198; Begg’s Test = 0.2458, p = 0.190] and in the ALSFRS-R scores [Egger’s Test = 1.18, p = 0.239; Begg’s Test = 0.58, p = 0.559]). Additionally, in ***Fig.4(b)*** the risk of bias analysis indicated that most clinical trials considered in the study were low-risk, with low concerns about missing outcomes and deviations from the intended interventions. Conclusively, both the parameters substantiated the reliance of the findings in the meta-analysis.

**Fig.4.**
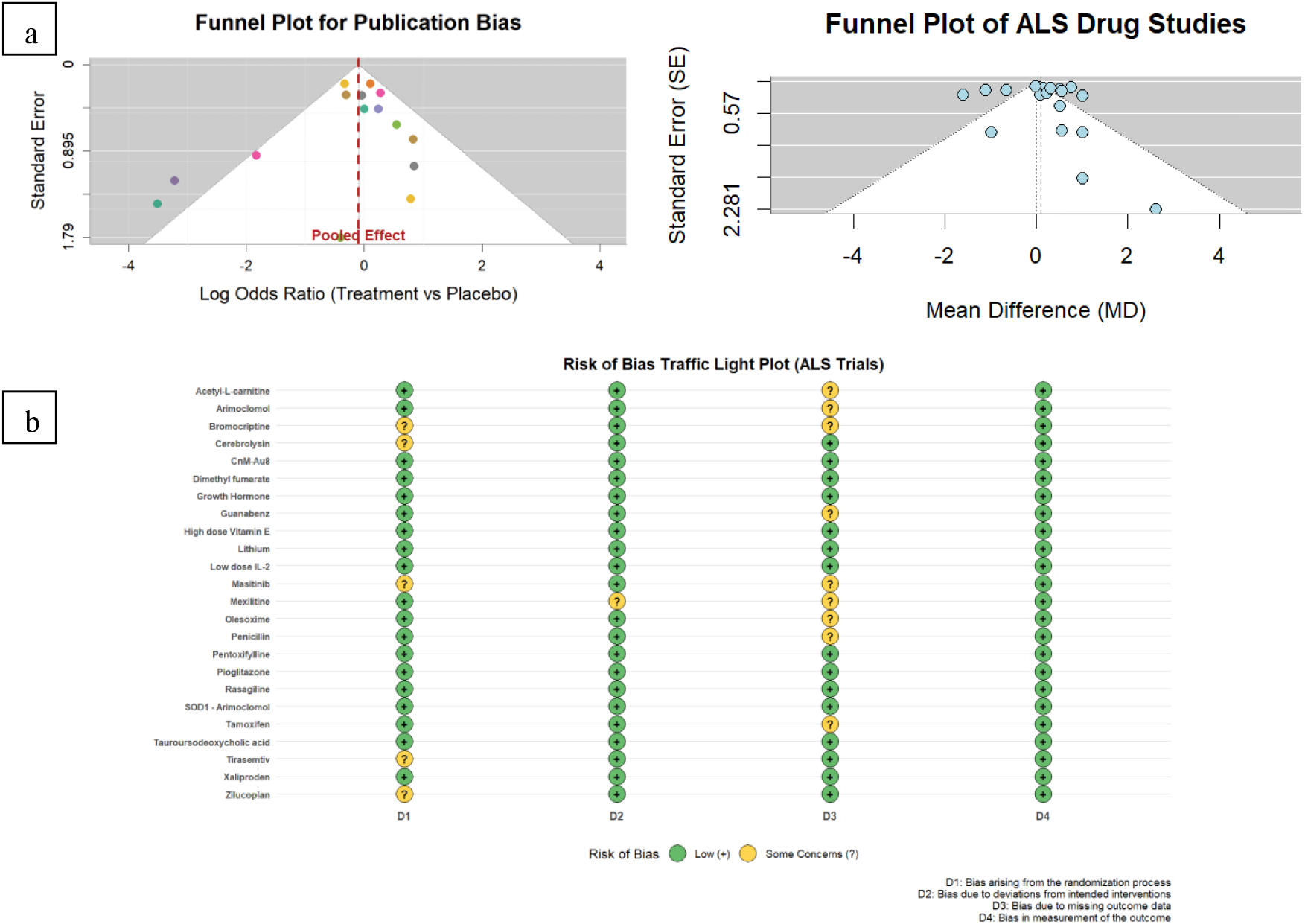
Evaluation of Publication Bias and Risk of Bias. ***(a)Funnel plot assessing publication bias in meta-analysis of Treatment vs. Placebo.*** The plot displays the log odds ratios (logOR) of individual studies (coloured dots) on the x-axis against their standard errors on the y-axis. The vertical dashed line represents the pooled effect estimate from the meta-analysis. The shaded funnel area indicates the expected 95% confidence region where studies would fall in the absence of publication bias and with sampling variation alone. While most studies lie within this region, a slight asymmetry is observed with fewer small studies reporting negative or null effects. This asymmetry suggests the potential for publication bias or small-study effects; however, corresponding statistical tests (Egger’s and Begg’s) did not detect significant bias. Overall, the meta-analytic estimate shows no clear effect favouring treatment or placebo. ***Funnel plot assessing publication bias in ALS drug studies***. The funnel plot depicts the relationship between study precision (standard error, SE) and mean difference (MD) across included studies evaluating pharmacological interventions in amyotrophic lateral sclerosis (ALS). Each circle represents an individual study, with the vertical dashed line indicating the pooled mean effect and the diagonal dotted lines showing the expected 95% confidence limits. The distribution of studies appears symmetrical, suggesting no apparent small-study or reporting bias. This visual interpretation is supported by Egger’s regression test (z = 1.1771, p = 0.2392) and Begg’s rank correlation test (z = 0.58, p = 0.5586), both indicating no significant evidence of publication bias. (***b)*** This figure summarizes the domain-specific risk of bias assessments for randomized controlled trials of therapeutic interventions in amyotrophic lateral sclerosis (ALS). Each circle represents one domain per study, colored according to the judged risk: green for *low risk of bias*, yellow for *some concerns*, and red for *high risk of bias*. Symbols within circles indicate the judgment level — “+” for low risk, “?” for some concerns, and “×” for high risk. Gridlines separate studies and domains for visual clarity.

### Two-dimensional Uniform Manifold Approximation and Projection of trial-level efficacy

As in ***Fig.5*** Unsupervised UMAP clustering of trial-level efficacy data identified four distinct drug behavior patterns in ALS. Cluster 1 included agents with robust or statistically significant slowing of functional decline (e.g., Rasagiline, CnM-Au8, Masitinib), while Cluster 3 comprised treatments associated with faster disease progression (e.g., Dimethyl fumarate, Arimoclomol). Clusters 2 and 4 demonstrated heterogeneous or variable responses. These findings highlight increased therapeutic benefit among mitochondrial and neuronal modulators.

**Fig.5.**
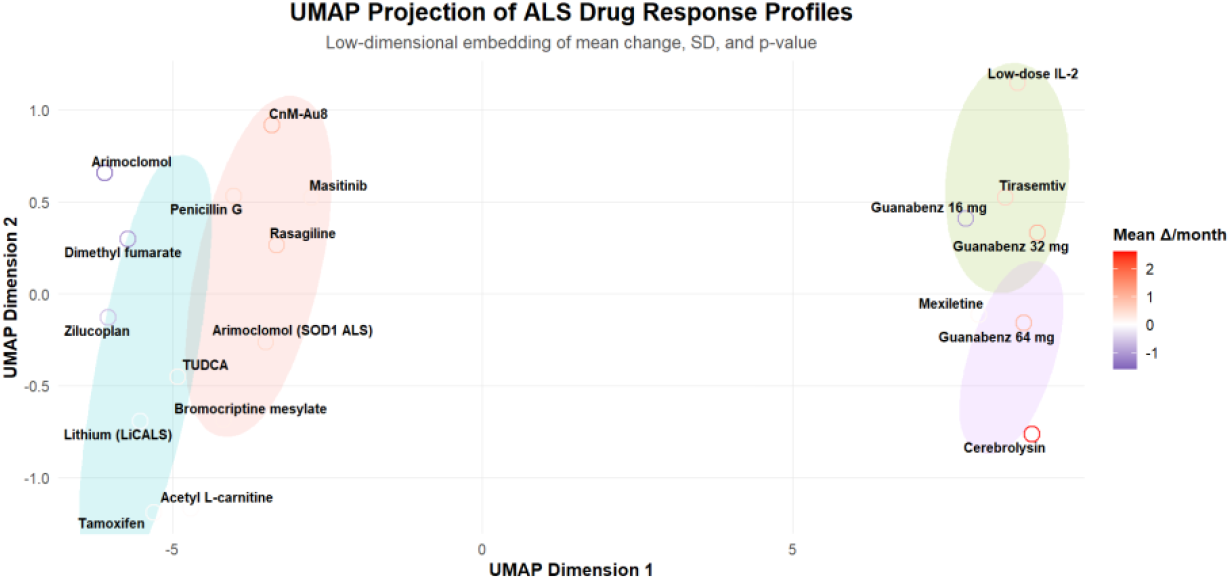
UMAP projection of ALS drug response profiles. A two-dimensional Uniform Manifold Approximation and Projection (UMAP) embedding of trial-level efficacy data revealed four distinct clusters of ALS drug response patterns based on mean functional change, standard deviation, and p-value. Cluster 1 (red) included agents demonstrating robust or statistically significant slowing of functional decline (e.g., rasagiline, CnM-Au8, masitinib). Cluster 3 (blue) comprises compounds associated with faster disease progression (e.g., dimethyl fumarate, arimoclomol). Clusters 2 (green) and 4 (purple) exhibited heterogeneous or variable responses, including drugs such as low-dose IL-2, trazemtiv, and guanabenz. These findings suggest convergent therapeutic benefit among agents targeting neuronal energy metabolism or mitochondrial stability, underscoring shared mechanistic pathways in ALS neuroprotection.

### Adverse events were observed across therapeutic interventions in both the treatment and placebo groups

The odds ratios for adverse events among the drug candidates showed no statistically significant difference between groups. While the overall dataset is not indicative of any marked pattern of consistency in adverse events between co-therapy and monotherapy, it does suggest the need for larger trials for examining the consistency. Refer to ***Fig.6(a) and (b)***.

**Fig.6.**
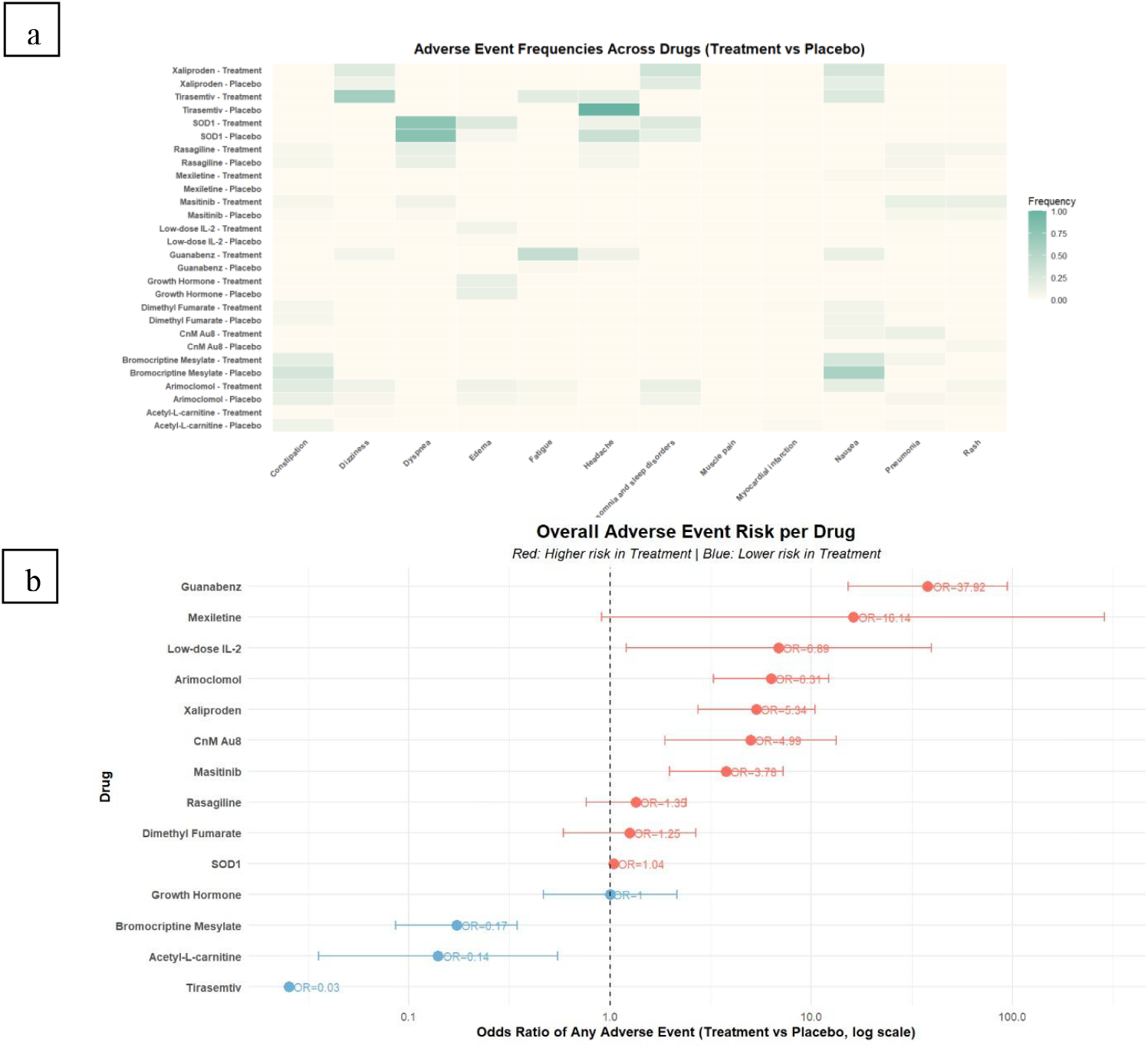
***(a) Heatmap of adverse event frequencies across pharmacological interventions in ALS clinical trials (co-therapy vs. monotherapy).*** The heatmap illustrates the relative frequencies of specific adverse events reported in randomized controlled trials comparing pharmacological co-therapies with monotherapy in amyotrophic lateral sclerosis (ALS). Rows represent individual drug–arm combinations (co-therapy or monotherapy), and columns correspond to distinct categories of adverse events. Colour intensity reflects the normalized frequency of each event, with darker shading indicating higher occurrence. ***(b) Overall adverse event risk associated with pharmacological cotherapies in ALS clinical trials (co-therapy vs. monotherapy)***. The forest plot displays odds ratios (OR) and 95% confidence intervals (CI) for the risk of experiencing any adverse event across amyotrophic lateral sclerosis (ALS) trials comparing pharmacological co-therapies with monotherapy. Each point represents the estimated OR for a given drug, plotted on a logarithmic scale. OR > 1 (shown in red) indicates a higher risk of adverse events with co-therapy relative to monotherapy, whereas OR < 1 (shown in blue) reflects a lower risk with co-therapy. The vertical dashed line at OR = 1 denotes no difference in risk between the two treatment strategies.

### Demographics and Study Gap

Most studies had male-to-female ratios greater than 1. A single study had equal male and female participants and one study had more female participants than women ***(refer to Fig.7(a))***. Additionally, a map of publications from 2004 to 2025 highlighting the temporal and geographical aspects was illustrated to decipher the gap in ALS research and the need for advancement in research ***(refer to Fig.7(b))***. The map showed a steady increase in investigations, hinting at more engagement in ALS therapeutics research over time. However, engagement was mostly confined to the USA and Europe, which showed a lack of trials outside of the area. This indicates a gap in access to trials for ALS patients outside the areas as well as a lack of representation of participants. Together, they provide information about the skewness in the recruitment of both sexes and robust engagement in research in ALS therapies across the globe.

**Fig.7.**
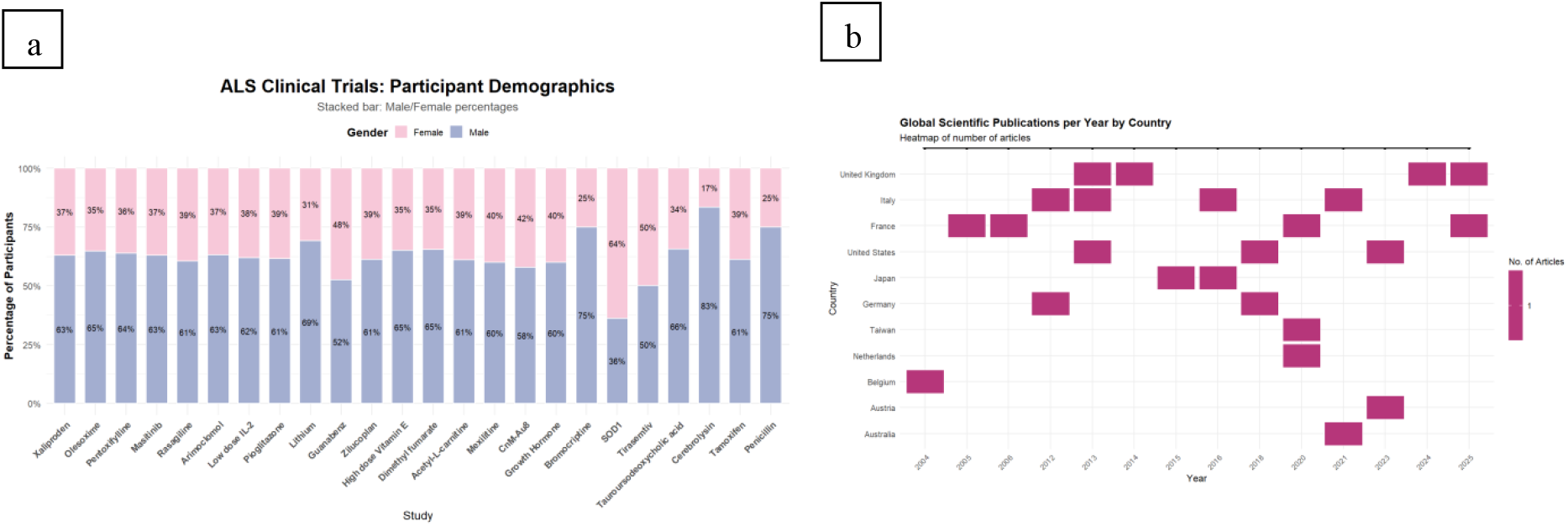
Demographic analysis of the RCT data. **(a)** *Gender distribution across ALS clinical trials*. Stacked bar plot showing the proportion of male and female participants enrolled in amyotrophic lateral sclerosis (ALS) clinical trials. The percentage of male participants (blue) consistently exceeded that of female participants (pink) across nearly all studies. **(b)** *Global scientific publications on ALS drug studies by country and year*. Heatmap illustrates the temporal and geographical distribution of ALS-related drug publications from 2004 to 2025. Each coloured cell represents the presence of one or more scientific articles published in a given year by researchers from the indicated country.

## Discussion

Despite newer approved therapies like edaravone and sodium phenylbutyrate/taurursodiol, riluzole remains a standard in ALS care. Our analyses show no significant difference between the safety and efficacy of riluzole monotherapy and co-therapy in ALS treatment. One of the primary features of riluzole treatment of ALS is prolonging life, even if by a few months; this was not augmented by any therapeutic agents. This makes monotherapy a better treatment strategy than any co-therapeutic drugs included in the study.

Current research in ALS treatment targets various common cellular pathways involved in disease progression. The drugs included in the study are modulators of neurotransmission, mitochondrial/oxidative stress, metabolic and signaling molecules, ion and voltage channels, and inflammation, reflecting that variation. A Unified Manifold Approximation and Projection based on survival, efficacy, and safety data clustered drugs based on performance as co-therapeutic agents with riluzole. The cluster with the most positive effect in ALS treatment (Cluster 1) comprises Rasagiline, CnM-Au8, Masitinib, Arimoclomol, TUDCA, and Bromocriptine. The cluster is primarily a combination of neuroimmune modulators and mitochondrial or oxidative stress modulators. Riluzole itself is a glutamate inhibitor that decreases neuronal excitability. This suggests that a combination of drugs targeting neural and mitochondrial/oxidative stress may be a rewarding direction for prospective research.

### Limitations

A limitation of clinical trials in ALS research is that the primary analysis of disease progression is done using ALSFRS-R score, which measures progression symptomatically. It remains the gold standard for clinical reporting in ALS. The lack of common use of established biomarkers decreases the sensitivity of efficacy assessment and prevents analysis of their impact on early ALS presentation. In some studies, biomarker profiling was performed using regulatory T-cell (Treg) counts, CSF-phosphorylated neurofilament heavy chain (CSF-pNFH), plasma and CSF chemokine ligand 2 (CCL2) concentrations, and urinary p75ECD protein levels. Despite recent efforts towards the establishment of minimally- or non-invasive biomarkers for the measurement of ALS progression, it is rare to see them reported in clinical trials.

There have been reports of different presentation, onset, and progression of ALS based on gender, yet the male/female ratio of participants tended to be greater than one. This may be because ALS tends to occur more frequently in men than women. However, women tend to present later than men, and are more likely to have bulbar onset and faster progression. Consistent overrepresentation of men in the participant pool can mask drugs that may be more effective against how ALS progresses in women. This may also skew safety and efficacy results for different types of ALS. Subgroup analysis and an equitable gender ratio can correct for this bias.

Efforts were made to include studies from around the world, but the selected studies (including multicenter studies) tended to preferentially include participants from countries with predominantly European-descended populations. This results in an overrepresentation of Caucasian individuals in ALS critical trials. Given the genetic basis of ALS, it is imperative to account for genetic variations that arise from ethnic differences when measuring the progression of ALS and drug efficacy against it.

In 2025, ALS remains a highly prevalent and largely unsolved challenge to public health. The complex and rapidly degenerative nature of the disease demands more robust and directed efforts focused on both diagnosis and management of the condition. The glaring gaps in ALS care and research, like the lack of biomarker-based progression data, impact not only our understanding of the disease but also our ability to best care for patients afflicted with it.

## Data Availability

All data produced in the present work are contained in the manuscript

## Acknowledgment

**HSR, JSB, SG, ND** acknowledge the Department of Biotechnology, Government of India for providing them the fellowship.

## CRediT authorship contribution statement

**HSR:** Conceptualization, Data Curation, Formal Analysis, Investigation, Methodology, Validation, Writing – Original draft, Writing – Review & Editing. **JSB:** Data Curation, Investigation, Methodology, Validation, Writing – Original draft. **SG:** Data Curation, Investigation, Methodology, Validation, Writing – Original draft. **ND:** Data Curation, Investigation, Methodology, Validation, Writing – Original draft. **SaK:** Investigation. **HaSR:** Investigation. **DB:** Methodology, Writing – Review & Editing, Software, Supervision, Project Administration. **SK: :**Conceptualization, Methodology, Writing – Review & Editing, Software, Supervision, Project Administration

## Data availability

Data generated during the study is part of the manuscript.

